# Gut colonization of *Enterococcus* species is associated with COVID-19 disease in Uganda

**DOI:** 10.1101/2024.09.28.24314457

**Authors:** Carolina Agudelo, David Patrick Kateete, Emmanuel Nasinghe, Rogers Kamulegeya, Christopher Lubega, Monica M Mbabazi, Noah Baker, Kathryn Lin, Chang C. Liu, Arthur Shem Kasambula, Edgar Kigozi, Kevin Komakech, John Mukisa, Kassim Mulumba, Patricia Mwachan, Brenda Sharon Nakalanda, Gloria Patricia Nalubega, Julius Nsubuga, Diana Sitenda, Henry Ssenfuka, Giana Cirolia, Jeshua T. Gustafson, Ruohong Wang, Moses Luutu Nsubuga, Fahim Yiga, Sarah A. Stanley, Bernard Ssentalo Bagaya, Alison Elliott, Moses Joloba, Ashley R. Wolf

## Abstract

**Background:** Infection with the COVID-19-causing pathogen SARS-CoV-2 is associated with disruption in the human gut microbiome. The gut microbiome enables protection against diverse pathogens and exhibits dysbiosis during infectious and autoimmune disease. Studies based in the United States and China have found that severe COVID-19 cases have altered gut microbiome composition when compared to mild COVID-19 cases. We present the first study to investigate the gut microbiome composition of COVID-19 cases in a population from Sub-Saharan Africa. Given the impact of geography and cultural traditions on microbiome composition, it is important to investigate the microbiome globally and not draw broad conclusions from homogenous populations.

**Results:** We used stool samples in a Ugandan biobank collected from COVID-19 cases during 2020-2022. We profiled the gut microbiomes of 114 symptomatic individuals who tested positive for SARS-CoV-2 along with 76 household contacts who did not present any symptoms of COVID-19. The inclusion of healthy controls enables us to generate hypotheses about bacterial strains potentially related to susceptibility to COVID-19 disease, which is highly heterogeneous. Comparison of the COVID-19 patients and their household contacts revealed decreased alpha diversity and blooms of *Enterococcus* and *Eggerthella* in COVID-19 cases.

**Conclusions:** Our study finds that the microbiome of COVID-19 individuals is more likely to be disrupted, as indicated by decreased diversity and increased pathobiont levels. This is either a consequence of the disease or may indicate that certain microbiome states increase susceptibility to COVID-19 disease. Our findings enable comparison with cohorts previously published in the Global North, as well as support new hypotheses about the interaction between the gut microbiome and SARS-CoV-2 infection.

## Background

Infection with Severe Acute Respiratory Coronavirus 2 (SARS-CoV-2) has extremely variable presentation, ranging from asymptomatic to fatal^1^. The resulting disease COVID-19 arose in December 2019 and has caused a global pandemic. The World Health Organization reports 775 million SARS-CoV-2 infections and 7 million deaths worldwide, including 9.6 million infections and 175 thousand deaths in the African continent^2^. SARS-CoV-2 infection results in highly heterogeneous disease, marked by fever, cough, fatigue, and other flu-like clinical symptoms. Underlying conditions increasing risk for severe disease include chronic lung problems, heart disease, diabetes, obesity, advanced age and pregnancy^1^. However, even young, healthy individuals have died from COVID-19, and what drives disease heterogeneity remains largely unexplained. Sub-Saharan Africa experienced a lower disease burden of COVID-19 than expected for reasons that remain underexplored^3,4^, although the microbiome is hypothesized to play a role.

The mammalian gut microbiome influences susceptibility to diverse pathogens and varies substantially across geographies. The protection of the gut microbiota against pathogens can occur directly, through bacterial competition in the gut, and indirectly, through modulation of the host immune system. In mouse models, the gut microbiome impacts transmission and pathogenesis of lung pathogens including *Klebsiella spp*. and *Burkholderia thailandensis*^5,6^. Gut helminths have also been shown to alter the microbiome and immune signaling to impact respiratory syncytial virus (RSV)^7,8^.

The gut microbiome of COVID-19 patients has been described previously, but published studies have significant limitations in the numbers of patients evaluated and are primarily restricted to individuals in the United States, Europe, and China^9–11^. Many of these studies lack samples from asymptomatic controls and none include individuals living in sub-Saharan Africa. These descriptive studies have been powerful to describe how the gut microbiome changes during the course of COVID-19 and also how it compares between severe and less severe COVID-19 cases. However, this research is limited in its ability to generate hypotheses about the role of the gut microbiome in protecting against SARS-CoV-2 infection or COVID-19 disease severity.

A systematic review^12^ identified 22 articles exploring 16S rRNA gene amplicon sequencing or metagenomic sequencing of patients with COVID-19. These studies identified alterations of diverse bacterial genera in COVID-19 patients including: depletion of *Ruminococcus, Alistipes, Eubacterium, Bifidobacterium, Faecalibacterium, Roseburia, Fusicathenibacter*, and *Blautia* and enrichment of *Eggerthella, Bacteroides, Actinomyces, Clostridium, Streptococcus, Rothia*, and *Collinsella*. An increase in *Enterococcus* abundance was identified in severe COVID-19 cases in studies conducted at NYU Langone Health and Yale New Haven Hospital, the University of Chicago Medical Center, in Hong Kong, and at the Technical University Hospital of Munich^11,13–15^. The majority of these study sizes were small (less than 100 individuals), and just one was from the African continent (Egypt)^16^. Given the known heterogeneity of the gut microbiome across geographic regions, it is important to study the interaction of COVID-19 and the gut microbiome in diverse populations and locations.

There are various hypotheses as to why Uganda and other African countries had lower COVID-19 case fatality rates (CFR). The lower CFR in Uganda may be due to the relatively low population density of an aging population (median age ∼17 years according to the 2024 national census), when compared with high-income countries, for example, the United States with a median age of (median age ∼38 years)^17,18^. Countries with a higher burden of individuals who have survived cardiovascular disease also had a higher mortality rate, and underreporting may contribute^18^. The gut microbiome differs substantially between individuals in Uganda and those in previously studied contexts like the United States and China. Given the role of the microbiome in immune maturation and protection from diverse infectious diseases, we sought to study the role of the gut microbiome in COVID-19 cases in Uganda.

Here we describe the gut microbiomes of individuals from Uganda with symptomatic COVID-19 and a set of healthy asymptomatic controls, whose SARS-CoV-2 infection status was not verified by PCR test, from similar geographic locations. We find that the microbiomes of COVID-19 cases are more dispersed than controls, and that a subset of individuals with COVID-19 have substantial *Enterococcal* blooms. Future investigations may be able to determine whether these microbiome disruptions impact the course of COVID-19 disease or recovery.

## Methods

### Sample collection

The study received ethical approval from the Mulago Hospital Research & Ethics Committee and the Uganda National Council for Science and Technology in June 2020 (approval #s MHREC1868 & HS869ES). We used stool samples from the protocol “Establishment of a Quality Assured COVID-19 Specimen Repository to Support Research in Diagnosis, Prevention and Management of SARS CoV-2 in Uganda”^19^. The study recruited symptomatic, PCR-positive COVID-19 patients, and their household contacts in Uganda from June 2020 to December 2022. PCR-positive participants were enrolled from government-designated COVID-19 treatment centers around the country, with most study participants recruited in the capital city, Kampala. All study participants provided written informed consent before being included in the study.

Samples for SARS-CoV-2 PCR-positive patients were on average collected within four days of a positive PCR result. PCR-positive patients were further consented to recruit their household contacts, who were included as asymptomatic controls if they were free of COVID-19 symptoms (these individuals were not PCR tested due to regulations governing use of scarce testing supplies at the time). Blood (plasma, serum), saliva, urine, stool, and oropharyngeal/nasopharyngeal swabs were collected from each participant at baseline. Samples collected were processed and stored at the Integrated Biorepository of H3Africa Uganda (IBRH3AU). Aliquots of stool from 190 participants were used for this study.

### Clinical data

Data was collected using an electronic questionnaire as described in Kamulegeya et al.^19^. This questionnaire collected data on participant sociodemographic factors, occupation, marital status, education background, travel history, and current and past health conditions. In addition to the questionnaire, an interview was also administered by a trained nurse. Available metadata is compiled in Supplemental Table 1.

### DNA isolation and 16S rRNA gene amplicon library preparation

Genomic DNA (gDNA) was isolated using the DNeasy PowerLyzer PowerSoil Kit from Qiagen (Cat. No. / ID: 12855-50). Following the manufacturer’s protocol, one scoop of stool was taken from each stool aliquot for gDNA extraction. gDNA was quantified using a Nanodrop and diluted to 10 ng/l. 16S rRNA amplification was conducted using the KAPA HiFi Hotstart kit (Cat. No. 50-196-5215) and the primers targeting the V4 region as described by Gohl et al.^20^ (Supplemental Table 2). Cycling conditions were as follows: 95 °C for 5 min, followed by 25 cycles of 98 °C for 20 s, 55 °C for 15 s, and 72 °C for 1 min. Final extension for 10 min at 72 °C. A second PCR followed to add sample-specific indices and Illumina compatible flow cell adaptors (Supplemental Table 2). Cycle was as follows: 95 °C for 5 min; ten cycles of 98 °C for 20 s, 55 °C for 15 s, and 72 °C for 1 min; and a final extension at 72 °C for 10 min. A left-side selection (0.8X) using SPRI beads followed the PCR using the manufacturer’s protocol. The PCR product was quantified using a Qubit and pooled for sequencing on an Illumina MiSeq to generate 2×250 bp paired end reads.

### Microbiome data analysis

Each of the 126 samples included in analysis contained a minimum number of 1000 reads per sample (Supplemental Table 1). Demultiplexed reads were pooled from multiple sequencing runs. Reads were demultiplexed using standard Illumina software or the iu-demultiplex package^21^. The Quantitative Insights into Microbial Ecology 2 (Qiime2) pipeline was used for downstream analyses^22^. Potential chimeric reads were removed using consensus-based methods. Amplicon sequence variants (ASVs) were inferred using DADA2^23^ and a phylogenetic tree was built using MAFFT alignment^24,25^. Taxonomic assignment was performed using the SILVA138 database^26^. For alpha and beta diversity analyses, samples were rarefied to1000 reads per sample using the Qiime2 pipeline.

### Statistical analysis

Alpha diversity was calculated using Faith’s Phylogenetic Diversity (PD) and the Shannon Index, with between group significance determined using a Kruskal-Wallis test. Using the Qiime2 package, beta diversity was calculated using the Weighted UniFrac distance matrix; this matrix considers abundance of observed organisms. Weighted UniFrac was used to measure beta diversity. PERMANOVA tests were done to see if the distributions were different. Weighted UniFrac value was set as the dependent variable, while SARS-CoV2 infection, participant location, antibiotic use, *Enterococcus* spp. presence, age, and sex were used as the independent variables. PERMDISP was used to calculate whether the variances between two groups were significantly different. The metadata file and Qiime2 files were imported into R and merged into a single Phyloseq object. Analysis of composition of microbiomes with bias correction (ANCOM-BC)^27^ was performed in R using the “ANCOMBC”^27,28^ package. Bacterial taxa were determined to be differentially present based on an adjusted p-value of < 0.05. Plots were generated using the “ggplot2”^29^ package and base R functions and edited in Adobe Illustrator.

### Shotgun whole genome sequencing

The Qiime2 taxonomy feature table was used to determine which positive samples had *Enterococcus* species present. gDNA from these samples was used for shotgun whole genome sequencing using the published Hackflex protocol^30^. Samples were pooled and sequenced on an Illumina NovaSeq to obtain 2×150bp reads. Sequences were uploaded onto Chan Zuckerberg’s ID (CZ ID) online pipeline and reads per million calculated using the NT database^31^. Sequences were analyzed using the metagenomics Illumina pipeline 8.3.0 and the antimicrobial resistance pipeline 1.3.2.

### USA metagenomes meta-analysis

A meta-analysis of six metagenomic inflammatory bowel disease (IBD) studies from the United States was performed to evaluate the presence of *Enterococcus* species in additional patient populations and healthy controls^32–37^ (see Supplemental Table 4). All data was retrieved from the Sequencing Read Archive as raw fastq files and processed collectively through the CZ ID metagenomic pipeline which maps short read sequencing files to NCBIs bacterial nucleotide database. Sequences were uploaded onto the CZ ID online pipeline and analyzed as described above. Prior to analysis, samples were filtered to remove any IBD subjects in remission (non active disease states) and any time points where subjects were undergoing a clinical intervention. Patients and controls in these studies were separated into antibiotic status based on whether antibiotics had been used within 3 months.

## Results

### Development of a biobank of COVID-19 cases and controls in Uganda

Participants were recruited from three COVID-19 treatment centers: Mulago National Referral Hospital, Entebbe Grade B Referral Hospital, and Masaka Regional Referral Hospital from June 2020 to December 2022 as described in Kamulegeya et al.^19^ Individuals who tested PCR positive for SARS-CoV-2 and their asymptomatic household members were recruited to the study. The resulting COVID-19 biobank managed by the Integrated Biorepository of H3Africa Uganda (IBRH3AU) contains blood (plasma, serum), saliva, urine, stool, and nasal swabs. Participants also completed an electronic questionnaire for demographic data. Asymptomatic control individuals were not PCR tested for SARS-CoV-2 due to the global shortage of tests at the time, so their infection status is unknown.

We extracted DNA and generated 16S rRNA gene amplicon sequencing libraries from 190 stool samples. 126 libraries passed quality checks and were used for subsequent analysis (Supplemental Table 1). From these filtered samples, 101 samples had demographic metadata that allowed for more specific analyses, such as geographic location, sex/gender, age, and antibiotic treatment at the time of sample collection (Figure 1, Table 1).

**Fig 1.**
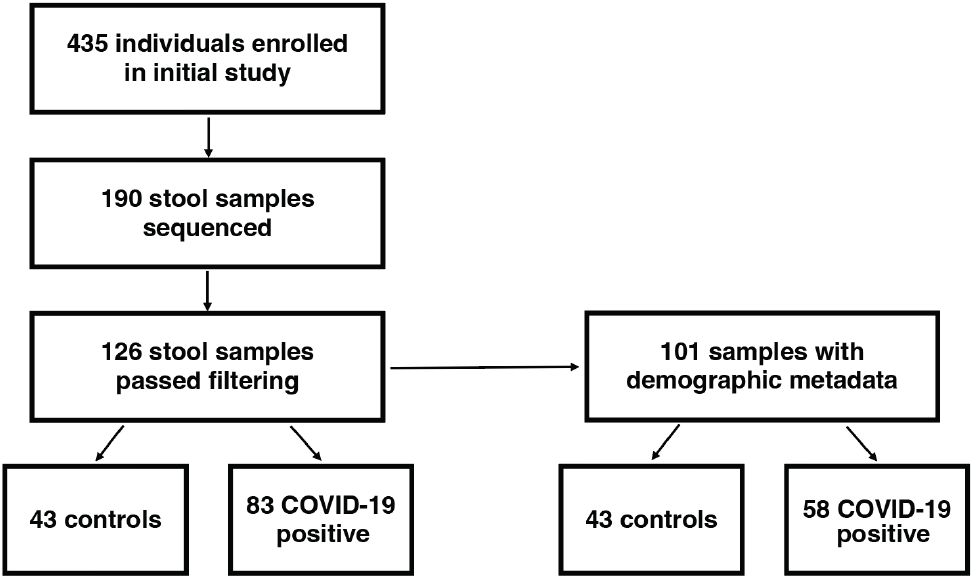
Flowchart of participants and analyzed stool samples.

**Table 1.** Sample characteristics with available metadata (N=101)

| Characteristic |  | n (%) or Median (IQR) |  |
| --- | --- | --- | --- |
| COVID-19 result |  | COVID-19 Positive | Asymptomatic control |
|  |  | 58 (57.4) | 43 (42.6) |
| Sex | Female | 21 (20.8) | 23 (22.8) |
|  | Male | 37 (36.6) | 19 (18.8) |
|  | Unknown | 0 (0.0) | 1 (1.0) |
| Age |  | 49 years (33-60) | 24.5 years (19-35.75) |
|  | <20 | 1 (1.0) | 12 (11.9) |
|  | 20-29 | 10 (9.9) | 16 (15.8) |
|  | 30-39 | 8 (7.9) | 6 (5.9) |
|  | 40-49 | 10 (9.9) | 5 (5.0) |
|  | 50-59 | 13 (12.9) | 2 (2.0) |
|  | 60-69 | 8 (7.9) | 0 (0.0) |
|  | >=70 | 8 (7.9) | 1 (1.0) |
|  | Unknown | 1 (1.0) | 1 (1.0) |
| Antibiotics | Yes | 30 (29.7) | 1 (1.0) |
|  | No | 28 (27.7) | 14 (13.9) |
|  | Unknown | 0 (0.0) | 28 (27.7) |

### Diversity of the gut microbiome during SARS-CoV-2 infection

We evaluated the composition of the gut microbiome of individuals positive for SARS-CoV-2 infection and asymptomatic controls using 16S rRNA gene amplicon sequencing of DNA isolated from stool samples. This allowed identification of most ASVs at the genus level. Based on the abundances of these ASVs, we calculated alpha diversity for each sample using a metric that includes phylogenetic information (Faith’s Phylogenetic Diversity (PD)) and a metric that is phylogeny independent (Shannon Index). We identified significantly lower alpha diversity in the gut microbiomes of COVID-19 cases compared to those of asymptomatic controls using both metrics, suggesting that COVID-19 cases have less complex microbiomes than controls (Fig. 2AB, Faith’s PD p*** < 0.0005, Shannon p**** < 0.000005, Kruskal-Wallis).

**Fig 2.**
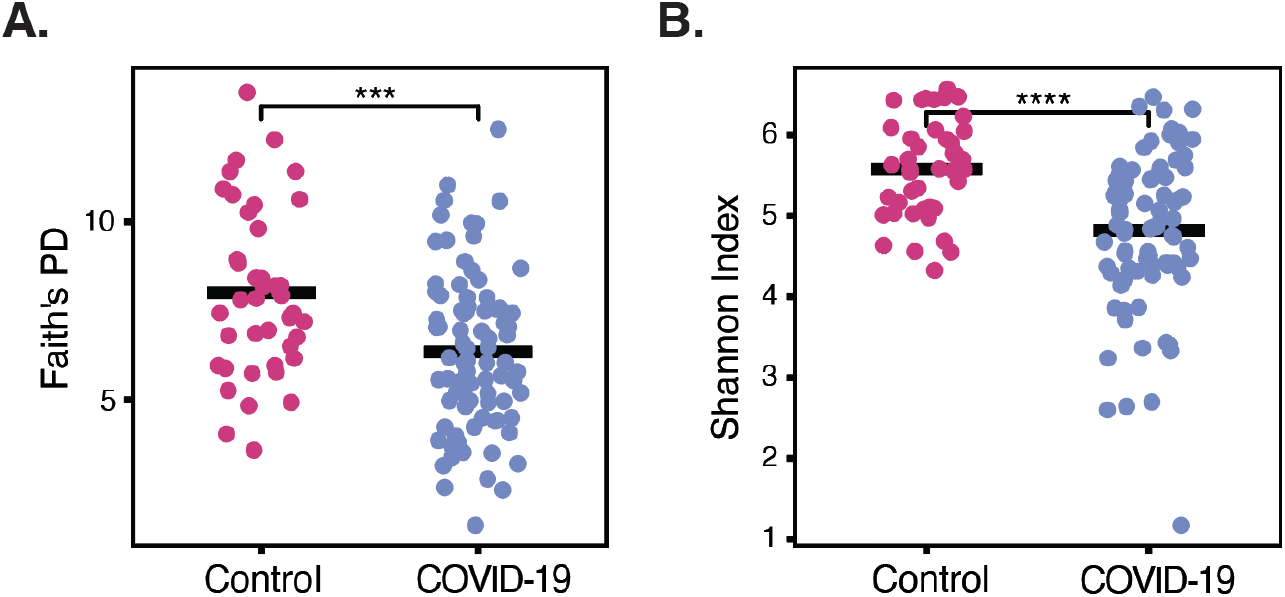
The gut microbiome of individuals with COVID-19 is less diverse. Alpha diversity is shown using **A** Faith’s Phylogenetic Diversity (PD) and **B** the Shannon Index (p*** < 0.01 Faith’s PD p*** < 0.001 Shannon, Kruakal-Wallia).

We next investigated the similarity of gut microbiomes across individuals using beta diversity. We used the Weighted UniFrac distance metric which takes into account both ASV abundance and taxonomy. The gut microbiomes of individuals infected with SARS-CoV-2 formed an overlapping, but distinct distribution by principal coordinate analysis when compared to asymptomatic controls (Fig. 3A, PERMANOVA p < 0.01). Microbiomes of asymptomatic individuals were more similar to each other, and those of infected individuals were significantly more dispersed (p < 0.01, PERMDISP). No differences were observed between the microbiomes of male and female subjects (Supplemental Figure 1A, p< 0.01). Age of subjects also did not appear to explain the increased dispersion (Supplemental Figure 1B).

**Fig 3.**
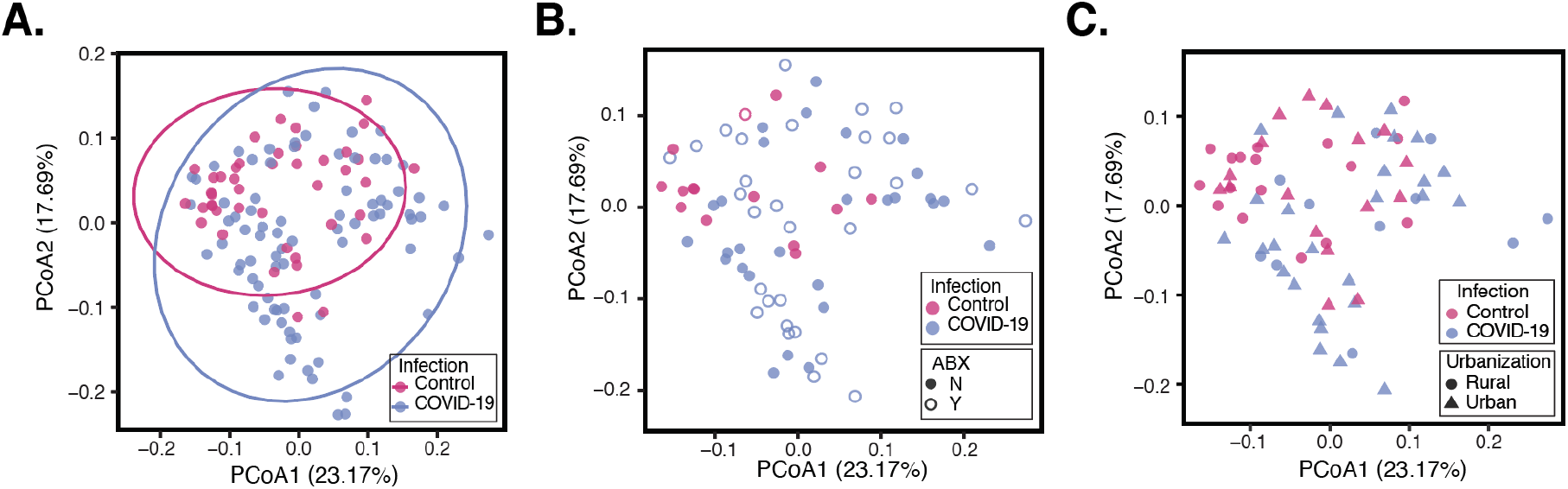
Beta-diversity analysis of the gut microbiomes of COVID-19 cases and asymptomatic controls. **A** Principal coordinate analysis (PCoA) calculated using the weighted UniFrac distance metric is shown for individuals who tested positive for COVID-19 and asymptomatic controls (p < 0.01, PERMANOVA; p <0.01, PERMDISP). The same PCoAis also plotted with point fill indicating recent antibiotic treatment at the time of sample collection (**B**, p = 0.577, PERMANOVA; p = 0.915, PERMDISP) or a symbol indicating whether the individual resides in a rural or urban area (**C**, p = 0.127, PERMANOVA; p = 0.515, PERMDISP).

We overlaid clinical and geographic data on the PCoA plot to see if these groupings drove the differences observed. We first wanted to test if antibiotic exposure explained the increased dispersion in COVID-19 cases. Comparing the distribution and dispersion of microbiomes from SARS-CoV-2 positive individuals with and without antibiotic treatment did not yield any significant differences (Fig. 3B, p = 0.577, PERMANOVA; p = 0.915, PERMDISP). We next tested the role of geography in microbiome composition. Both controls and COVID-19 cases were sampled from geographic regions around Uganda. Samples from individuals in the same district did not cluster with each other (Supplement Figure 1C). Districts were classified into major cities (urban) or rural (all other areas) based on country data^38^. Using these criteria, gut microbiome beta diversity also did not differ significantly based on district urbanization (Figure 3C, p = 0.127, PERMANOVA; p = 0.515, PERMDISP). Given that these variables are not significantly related to dispersion between microbiome samples, we conclude that COVID-19 cases are correlated with increased dispersion independent of the recorded metadata.

### Taxa enriched in COVID-19 cases or controls

We next explored whether specific taxa are enriched in the microbiomes of COVID-19 cases or controls. Analysis of microbiome composition with bias correction (ANCOM-BC)^27^ identified genera that were significantly enriched in asymptomatic controls. Species significantly enriched in controls with a log-2 fold change greater than 1.5 include *Lactobacillus, Clostridium sensu stricto 1*, genera of the *Pasteurellaceae* family, *Romboutsia, Agathobacter*, and *Akkermansia* (Figure 4A, Supplemental Figure 2). *Akkermansia* and *Romboutsia*, in particular, have been shown to be associated with beneficial outcomes in the context of inflammatory conditions such as diabetes and ulcerative colitis^39–41^.

**Fig 4.**
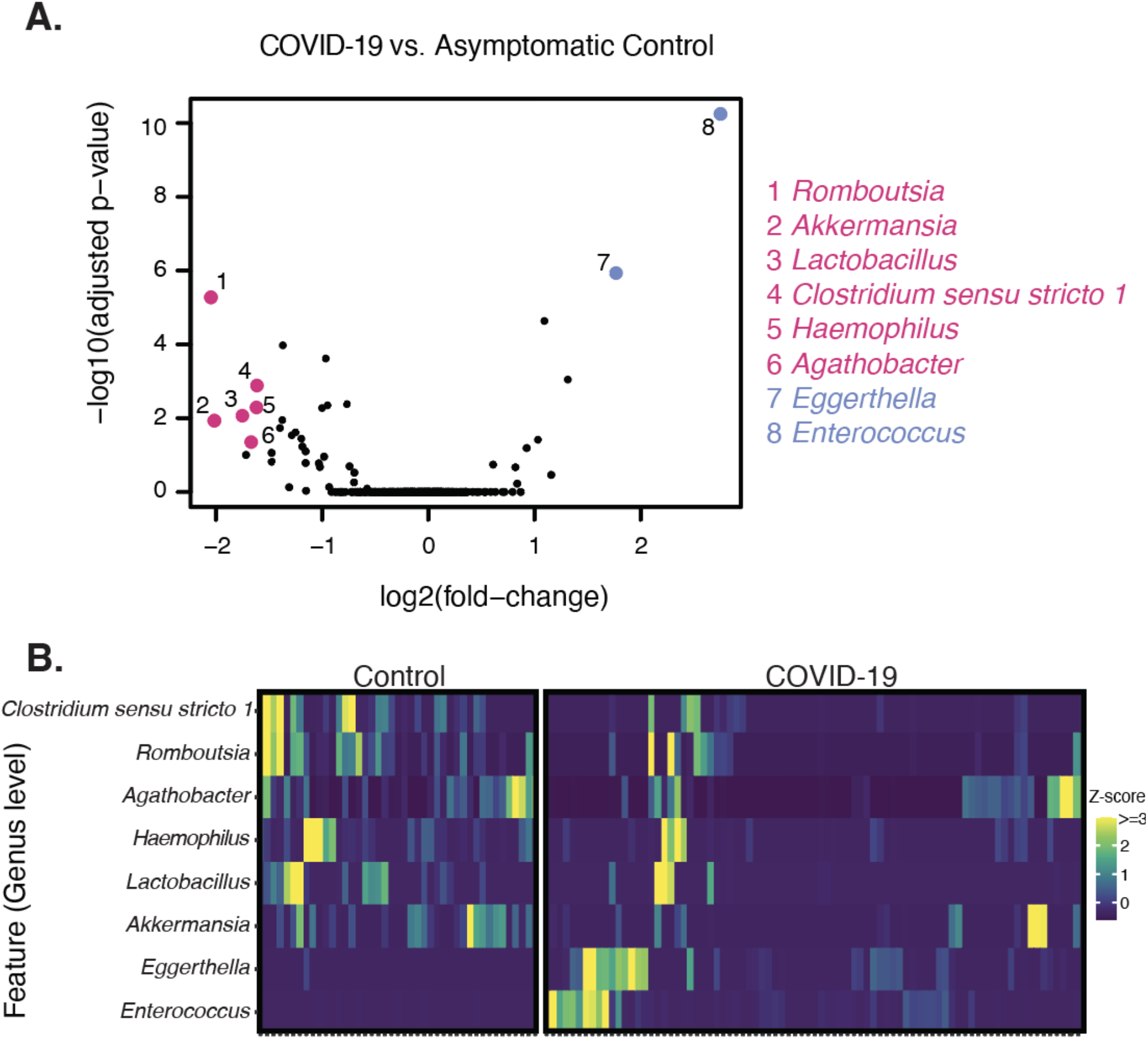
Genera differentially abundant between COVID-19 cases and controls. **A** The log2 fold change of each genera is plotted versus the -log10(Adj p-value) as calculated by analysis of composition of microbiomes with bias correction (ANCOM-BC) (highlighted species have a absolute log2-fold change greater than 1.5 and a p-value less than 0.05). **B** Relative abundance for each taxa identified in A was plotted as a z-score.

The gut microbiomes of COVID-19 cases were strikingly enriched for *Eggerthella* and *Enterococcus* (1.76-fold and 2.75-fold respectively, adjusted p-value < 0.05) (Figure 4B). *Enterococcus* species were completely absent from asymptomatic controls, whereas *Enterococcus* was found in 56.6 percent of COVID-19 cases. The range of *Enterococcus* relative abundance in the COVID-19 cases ranged from 0-85% (Figure 5A). *Eggerthella* species were present in only 2% of asymptomatic individuals, whereas *Eggerthella* was found in 43.4 percent of COVID-19 cases. *Eggerthella* relative abundance in COVID-19 cases ranged from 0-4% (Figure 5B).

**Fig 5.**
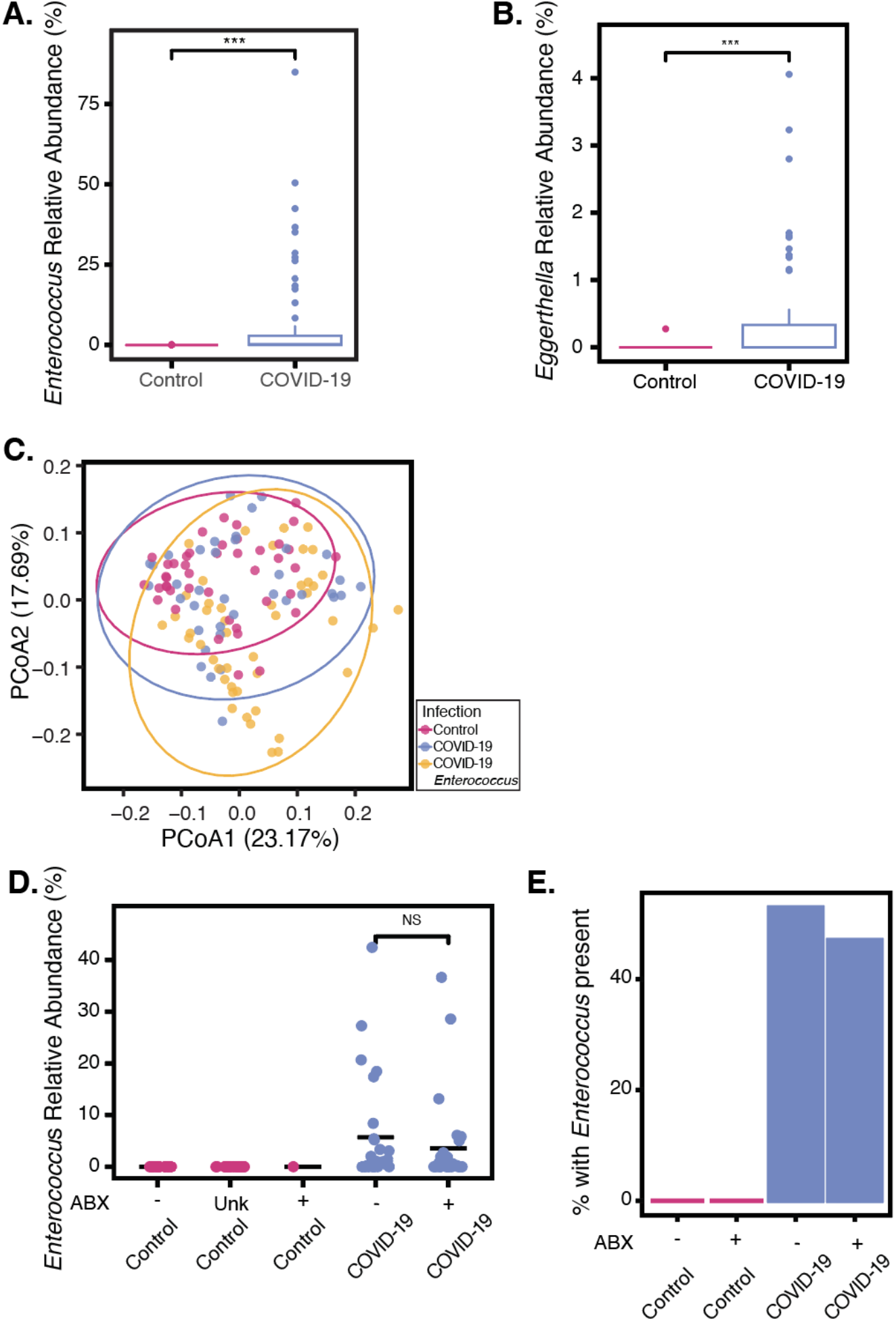
*Enterococcus* species were highly abundant in COVID-19 cases. The relative abundance of **A** *Enterococcus* species (p * * * < 0.0005, Kruskal-Wallis) and **B** *Eggerthella* species (p * * * < 0.0005, Kruskal-Wallis) in positive COVID-19 individuals and asymptomatic individuals. **C** PCoA identifying positive COVID-19 individuals with *Enterococcus* present in their gut microbiome (p < 0.01, PERMANOVA; p = 0.639, PERMDISP). **D** *Enterococcus* relative abundance in COVID-19 cases and controls based on antibiotic treatment status (ABX= antibiotic treated, Unk = Unknown, not significant -NS, Kruskal-Wallis). **E** The percent of individuals with *Enterococcus* detected at greater than 0.1 % in their gut microbiome is shown for asymptomatic controls and COVID-19 positive individuals.

We tested if *Enterococcus* presence drove the observed heterogeneity in beta diversity shown in Fig. 3A. We plotted the beta diversity PCoA and labeled the presence/absence of *Enterococcus* (Figure 5C). The microbiomes of COVID-19 cases that had *Enterococcus* did not cluster together, and formed a distinct distribution to the microbiomes of COVID-19 cases with no *Enterococcus* (p < 0.01, PERMANOVA). Microbiomes with *Enterococcus* did not cluster together and were quite dispersed. While this suggests that *Enterococcus* presence contributes to the broader dispersion seen in the microbiomes of COVID-19 cases, it was not sufficient to explain this increased dispersion. *Enterococcus* positive microbiomes were not significantly more dispersed than COVID-19 positive microbiomes without *Enterococcus* (p = 0.639, PERMDISP).

### Exploring the role of *Enterococcus* species in COVID-19

Antibiotic treatment has been previously shown to cause blooms in *Enterococcus* abundance in hospitalized patients with diarrhea in Vietnam or cancer in the United States^42,43^. Available metadata on antibiotic use allowed for us to test this hypothesis in the context of COVID-19 and Uganda. Individuals positive for SARS-CoV-2 were compared based on whether or not they received antibiotics at the time of sample collection. The relative abundance was also not significantly different based on antibiotic treatment within COVID-19 cases (Fig 5D, p = 0.626, Kruskal-Wallis test) suggesting that an increased risk of *Enterococcus* bloom in COVID-19 cases is not due to increased antibiotic treatment alone (16.8 % of COVID-19 cases without antibiotics contained *Enterococcus* vs. 13.9% of cases exposed to antibiotics, Fig 5E). Reported antibiotic exposure also did not result in significant changes in alpha diversity among COVID-19 positive individuals (Supplemental Figure 3AB). Interestingly, the vast majority of *Enterococcus* positive microbiomes were sampled from individuals at Mulago hospital (Supplemental Figure 4).

To determine the *Enterococcus* species contributing to these blooms, we performed shotgun metagenomic sequencing on five *Enterococcus*-containing samples. These samples were identified as Enterococcus-containing via the 16S rRNA amplicon sequencing. The metagenomes of the *Enterococcus*-containing samples were analyzed using the Chan Zuckerberg Infectious Disease (CZ ID) online platform^31^. These samples identified that the predominant species contributing to these blooms was *E. faecium. E. faecium* was identified in five of five sequenced samples (Figure 6A). *E. faecalis* was also identified in all samples sequenced at a lower percentage (Figure 6B). The antimicrobial resistance pipeline identified aminoglycoside, tetracycline, macrolide, streptogramin, lincosamide, and fluoroquinolone resistance reads associated with *E. faecium* (Supplement Table 3). Macrolide, fluoroquinolone, diaminopyrimidine, lincosamide, and pleuromutilin resistance reads were associated with *E. faecalis* (Supplement Table 3).

**Fig 6.**
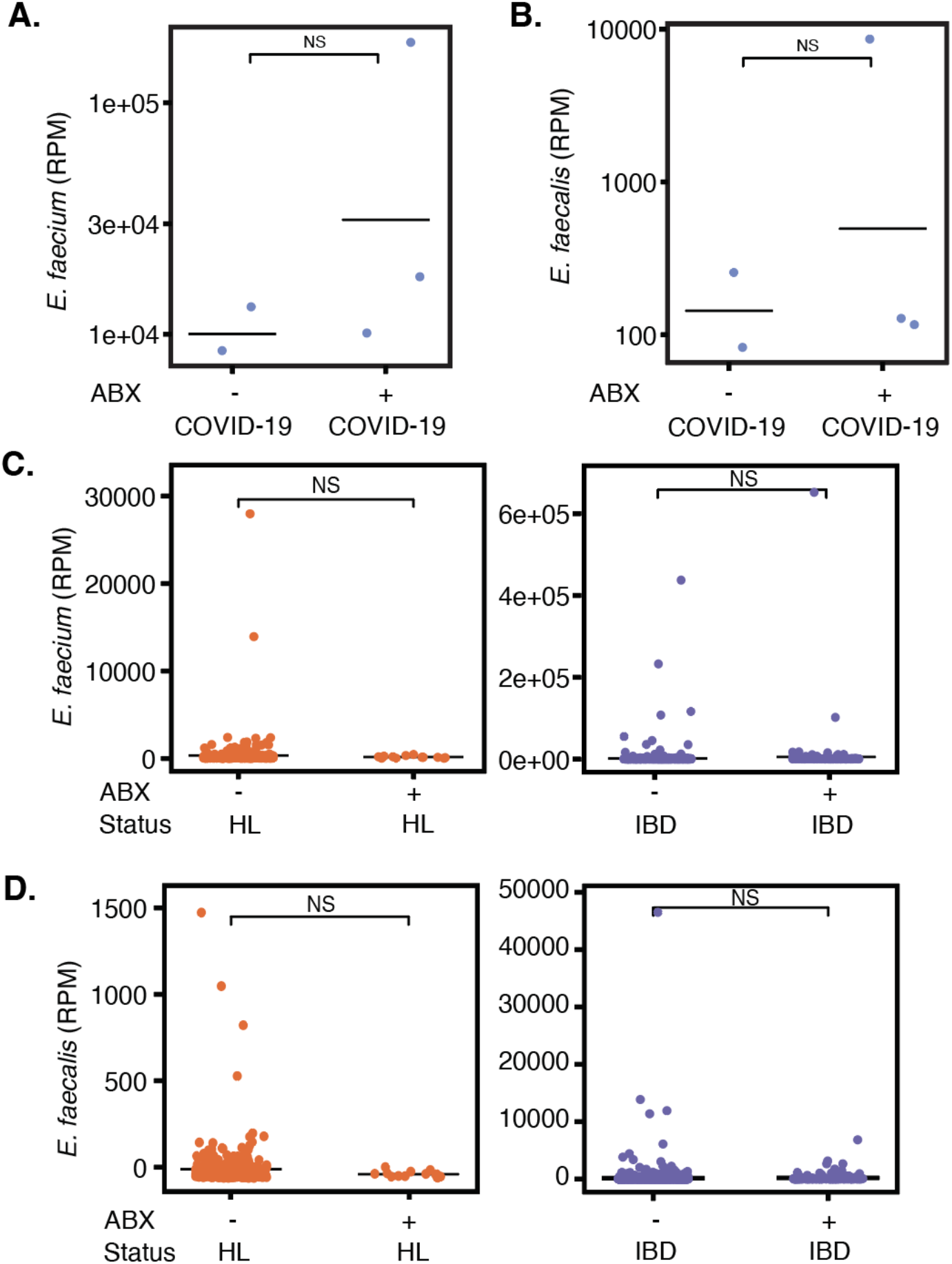
The role of antibiotic treatment on the abundance of *Enterococcus* species in different disease states. The reads per million (RPM) of **A** *Enterococcus faecium* (p = 0.25, Kruskal-Wallis) and **B** *Enterococcus faecalis* (p = 0.25, Kruskal-Wallis) in positive COVID-19 individuals with and without antibiotic treatment. A meta-analysis of USA studies with IBD patients and healthy controls was used to plot **C** *Enterococcus faecium* (p = 1, Wilcoxon test) and **D** *Enterococcus faecalis* (p = 0.92, Wilcoxon test) (ABX= antibiotic treated, HL = healthy, IBD = irritable bowel disease, not significant - NS, Kruskal-Wallis).

We next sought to understand the distribution of *Enteroccoccus* in individuals outside Uganda to compare to what we observed there. We re-analyzed gut metagenome data from six studies from the United States including healthy controls and patients with inflammatory bowel disease to identify levels of *Enterococcus* species^32–37^ (Supplemental Table 4). We found that *E. faecium* and *E. faecalis* were found at a low level in some healthy controls (less than 2500 reads per million), and that antibiotic exposure did not influence this. In fact, there was a trend toward antibiotic exposure suppressing *Enterococcus* (Figure 6CD). *Enterococcus* was more prevalent in individuals with inflammatory bowel disease, although again antibiotic history did not influence the abundance significantly (Figure 6CD, not significant, Wilcoxon test). Thus *Enterococcus* presence, like COVID-19 disease severity, may be heavily influenced by underlying medical conditions and geography.

## Discussion

During the SARS-CoV-2 pandemic, researchers reported on gut microbiome dysbiosis in cohorts of hospitalized COVID-19 patients located in the United States, Europe, and China^9– 11,13–15^. These studies identified bacterial taxa correlated with COVID-19 disease or its severity, but specific interactions and mechanisms were not identified. Given that the gut microbiome varies substantially across geographic and cultural contexts, it is important to also explore the relationship between the gut microbiome and COVID-19 in populations in the Global South, including Africa.

In this study, we found that Ugandan individuals who tested positive for SARS-CoV-2 had a less diverse gut microbiome compared to those of asymptomatic controls and increased dispersion. Decreased alpha-diversity has been observed in cohorts of diverse disease states including inflammatory bowel disease, flu, Parkinson’s disease, and multiple sclerosis^44–47^. Gu et al. also found lower diversity in COVID-19 cases compared to healthy controls in a Chinese study^9^. How microbial diversity decreases as an effect of infection or disease is still poorly understood. Our study includes asymptomatic individuals as controls for a healthy gut microbiome. In contrast many prior studies only compared COVID-19 patients at different levels of severity. Additionally, this study, to our knowledge, is the first COVID-19 microbiome cross-sectional study in sub-Saharan Africa. We saw no differences in the microbiomes between individuals in urban and rural areas. This may be because all individuals were able to access medical treatment and thus were relatively nearby to urban areas compared to other studies^48^.

Asymptomatic individuals recruited from the same households as cases were used as a proxy for a healthy microbiome. However, their SARS-CoV-2 infection status was not verified by PCR test. The microbiomes of asymptomatic individuals were enriched with *Romboutsia* and *Akkermansia. Romboutsia* was also identified in healthy controls when compared to individuals with COVID-19 in a study by Gu et al.^9^ This suggests a protective and beneficial role for *Romboutsia* in the context of SARS-CoV-2. *Romboutsia* species have also been associated with improvements in endothelial function and changes in metabolic function in diet-induced obese rats^41^. *Akkermansia*, on the other hand, is known for both its beneficial^49,50^ and detrimental roles^51^ in human health. *Akkermansia muciniphila* has been associated with alleviating ulcerative colitis and reducing the incidence of diabetes in mouse studies by modulating inflammatory gene expression^39,40^. It has also been associated with protection from infectious agents like *Listeria monocytogenes*^52^. In contrast, Bernard-Raichon et al.^11^ showed in their mouse model of SARS-CoV-2 that infection led to dysbiosis in the gut led by *Akkermansia*. This suggests a context-dependent role for *Akkermansia* in COVID-19 infection and disease and requires further investigation.

Individuals who tested positive for SARS-CoV-2 had higher relative abundances of microbiome-resident pathobionts than their asymptomatic counterparts. Both *Enterococcus* and *Eggerthella* can cause or exacerbate disease opportunistically, and both were elevated in the microbiomes of COVID-19 cases. Strikingly, no asymptomatic individuals had any *Enterococcus* present in their guts and just 2% of asymptomatic individuals had *Eggerthella* present at any abundance. *Enterococcus* is a member of the intestinal flora that can cause hospital-acquired bloodstream infections and urinary tract infections^53^. It has also been associated with states of gut dysbiosis as an effect of antibiotic usage^54^, although here we do not find that antibiotic increases *Enterococcus* carriage in our COVID-19 study or an meta-analysis of individuals from the United States. Other similar studies^11–15^ saw enrichment of *Enterococcus* in severe COVID-19 cases compared to less severe cases. Elevated *Enterococcus* abundance may lead to increased susceptibility to disease or be a consequence of COVID-19 infection and related treatment. Experimental models or prospective studies are required to define the role of *Enterococcus* carriage.

*Eggerthella lenta* is a gut pathobiont associated with bacteremia^55^ and inflammatory diseases. *Eggerthella* is elevated in patients with IBD^56^, and rheumatoid arthritis (RA) mouse models suggest *E. lenta* promotes IBD by antigen-independent Th17 activation^57^. COVID-19 disease has previously been associated with increased *Eggerthella* abundance in a study with just 13 cases and five controls^12,58^. Our study expands this finding and validates it in a larger sample size. Interestingly, Th17 responses have been associated with severe COVID-19^59,60^. *Eggerthella* may have a role in increasing host susceptibility to COVID-19 disease, possibly through exacerbation of Th17 responses, but further studies are needed.

Activation of the immune system by SARS-CoV-2 may impact gut microbiota composition, or gut microbiota composition may impact the immune response to SARS-CoV-2 infection. SARS-CoV-2 is largely a pulmonary infection, although a significant fraction of patients also experience infection of the gastrointestinal tract. Gut T cell priming against SARS-CoV-2 infection in the gut could be influenced by the microbiome. These T cells may subsequently traffic to the lungs, contributing to severe manifestations of infection. Even in the absence of gastrointestinal infection in the gut, the propensity of T cells to traffic from one mucosal tissue to another means that T cells specific for gut antigens could in principle influence antigen specific responses to COVID-19 in the lungs^61^. Conversely, T cell responses primed in the lung could influence the composition of the gut microbiome by altering the inflammatory environment in the intestinal mucosa. COVID-19 infection is associated with the production of cytokines including type I interferon, IL-6, IL-17, IL-1, and TNF-ɑ^60,62–64^. Cytokines produced during infection have been shown to have an impact on the gut microbiome^65,66^. In addition, specific gut microbes can impact production of cytokines^67,68^ that may impact the host’s response to the virus.

While our study provides advances on previous reports by including controls, a larger subject pool, and an understudied geographic region, it does have some limitations imposed by implementation challenges associated with the pandemic. We do not know whether the asymptomatic controls were asymptomatic carriers of SARS-CoV-2 or not infected. Regardless, the microbiome changes observed are interesting whether they are associated with resistance to infection or disease presentation. Future studies may be able to delve into this in more detail. We also do not have clinical metadata for ∼20% of the individuals with sequencing data, but the 101 individuals we did have is larger than most published studies. For controls, antibiotic use was not reported. Although individuals were sampled as soon as possible after diagnosis, there was variable time between start of infection and sampling. Despite these limitations, our analysis demonstrates for the first time disruption of the gut microbiome in individuals with COVID-19 in Uganda.

## Conclusion

We have shown an association with increased microbiome variability and an enrichment of *Enterococcus* species in the microbiomes of COVID-19 cases compared to asymptomatic controls in Uganda. These disruptions may be a consequence of the disease or cause increased susceptibility to SARS-CoV-2 infection and/or COVID-19 disease presentation. Mechanistic experiments or prospective cohort studies are required to disentangle these possibilities. Understanding how the gut microbiome interacts with respiratory pathogens can better inform treatment strategies and potentially the development of prophylactic probiotics.

## Data Availability

Sequencing reads have been submitted to the NCBI Short Read Archive under Bioproject PRJNA1131762. A link to the data is available by request.

## Declarations

### Ethics approval and consent to participate

The study received ethical approval from the Mulago Hospital Research & Ethics Committee and the Uganda National Council for Science and Technology in June 2020 (approval #s MHREC1868 & HS869ES). All participants consented to sample storage and use of their samples and data in current and future studies related to SARS-CoV-2 infection according to the protocol “Establishment of a Quality Assured COVID-19 Specimen Repository to Support Research in Diagnosis, Prevention and Management of SARS CoV-2 in Uganda”^19^.

### Consent for publication

Not applicable

### Availability of data and materials

Sequencing reads are available through NCBI Short Read Archive under Bioproject PRJNA1131762. Data is available by request during review.

### Authors’ contributions

C.A., E.N., and A.W. wrote the main manuscript text, prepared figures, and generated and analyzed data. E.N., R.K., C.L., M.M. and N.B. were responsible for Biobank sample coordination, and metadata organization. C.A., E.N., K.L., C.C.L., A.S.K., E. K., K. K., J. M., K. M., P. M., B.S.N., G.P.N., J.N., D.S., and H. S. were involved in DNA extraction and preparation of sequencing libraries. J.T.G, R.W., and G.C. analyzed data related to the USA meta-analysis. M.L.N. and F.Y. provided sequencing and bioinformatics support. S.S., D.P.K., B.B., M.J. and A.W. conceptualized the study and provided feedback on analysis. D.P.K., B.B., M.J., and A.E. oversaw cohort development and sample collection. All authors reviewed the manuscript and provided feedback.

## Acknowledgements

We thank Celine Perier and Isabelle Charles for their help organizing the Alliance summer workshop. We additionally thank Jessica Hoisington-Lopez and MariaLynn Crosby at Washington University in Saint Louis for sequencing support.

