## Supplemental Materials for "Gut colonization of *Enterococcus* species is associated with COVID-19 disease in Uganda"

#### **Supplemental Materials** *for Agudelo, Kateete, Nasinghe, et al.*

##### Suppl. Table 1 See attached

##### Suppl. Table 2

**Supplemental Table 2.** Primer and adaptor sequences

PCR1 primers - amplifying V4 region of 16S rRNA

| | |
|---|---|
| V4_515F_Nextera | TCGTCGGCAGCGTCAGATGTGTATAAGAGACAGGTGCCAGCMGCCGCGGTAA |
| V4_806R_Nextera | GTCTCGTGGGCTCGGAGATGTGTATAAGAGACAGGGACTACHVGGGTWTCTAAT |

Illumina Index adaptors

| | |
|---|---|
| Forward index | AATGATACGGCGACCACCGAGATCTACACXXXXXXXXTCGTCGGCAGCGTC |
| Reverse index | CAAGCAGAAGACGGCATACGAGATXXXXXXXXGTCTCGTGGGCTCGG |

##### Suppl. Table 3

**Supplemental Table 3.** AMR genes identified in *Enterococcus* species

| Antimicrobial resistance gene | Drug class | Mechanism | Model | Reads Species |
|---|---|---|---|---|
| AAC(6')-Ii | aminoglycoside antibiotic | antibiotic inactivation | protein homolog | *Enterococcus* (chromosome) |
| dfrE | diaminopyrimidine antibiotic | antibiotic target protection | protein homolog | *Enterococcus faecalis* (chromosome) |
| efmA | fluoroquinolone antibiotic; macrolide antibiotic | antibiotic efflux | protein homolog | *Enterococcus faecium* (chromosome) |
| efrA | fluoroquinolone antibiotic; macrolide antibiotic; rifamycin antibiotic | antibiotic efflux | protein homolog | *Enterococcus faecalis* (chromosome) |
| efrB | fluoroquinolone antibiotic; macrolide antibiotic; rifamycin antibiotic | antibiotic efflux | protein homolog | *Enterococcus faecalis* (chromosome) |
| ErmB | lincosamide antibiotic; macrolide antibiotic; streptogramin antibiotic | antibiotic target protection | protein homolog | *Enterococcus faecalis* (chromosome or plasmid) |
| ErmT | lincosamide antibiotic; macrolide antibiotic; streptogramin antibiotic | antibiotic target protection | protein homolog | *Enterococcus faecium* (chromosome or plasmid) |
| lsaA | lincosamide antibiotic; pleuromutilin antibiotic; steptogramin antibiotic | antibiotic target protection | protein homolog | *Enterococcus* (chromosome); *Enterococcus faecalis* (chromosome) |
| msrC | macrolide antibiotic; streptogramin antibiotic | antibiotic target protection | protein homolog | *Enterococcus faecium* (chromosome) |
| tet(L) | tetracycline antibiotic | antibiotic efflux | protein homolog | *Enterococcus* (chromosome or plasmid); *Enterococcus faecium* (chromosome or plasmid) |
| tet(M) | tetracycline antibiotic | antibiotic target protection | protein homolog | *Enterococcus faecium* (chromosome or plasmid) |
| tet(U) | tetracycline antibiotic | antibiotic efflux | protein homolog | *Enterococcus faecium* (chromosome or plasmid) |

### Suppl. Table 4

**Supplemental Table 4.** USA irritable bowel disease (IBD) studies used for meta-analysis of *Enterococcus* abundance.

| Study name | Citation | Studies by Geography | Locartion | SRA |
|---|---|---|---|---|
| Gut microbiome structure and metabolic activity in inflammatory bowel disease | Franzosa, Eric A et al., 2019 | USA_PRISM_1 | USA | PRJNA400072 |
| Short Article Gut Microbiome Function Predicts Response to Anti-integrin Biologic Therapy in Inflammatory Bowel Diseases | Ananthakrishnan, Ashwin N et al., 2017 | USA_PRISM_2 | USA | PRJNA384246 |
| Gastrointestinal Surgery for Inflammatory Bowel Disease Persistently Lowers Microbiome and Metabolome Diversity | Fang, Xin et al., 2021 | USA_QIITA_11546 | USA | PRJEB38352 |
| Multi-omics analyses of the ulcerative colitis gut microbiome link Bacteroides vulgatus proteases with disease severity | Mills, Robert H et al., 2022 | USA_QIITA_11549 | USA | PRJEB42151 |
| Dietary manipulation of the gut microbiome in inflammatory bowel disease patients: Pilot study | Olendzki, Barbara et al., 2022 | USA_umass | USA | PRJNA642308 |
| Overrepresentation of Enterobacteriaceae and Escherichia coli is the major gut microbiome signature in Crohn's disease and ulcerative colitis; a comprehensive metagenomic analysis of IBDMDB datasets | Khorsand, Babak et al., 2022 | USA_hmbp | USA | PRJNA400072 |

#### Suppl. Fig 1

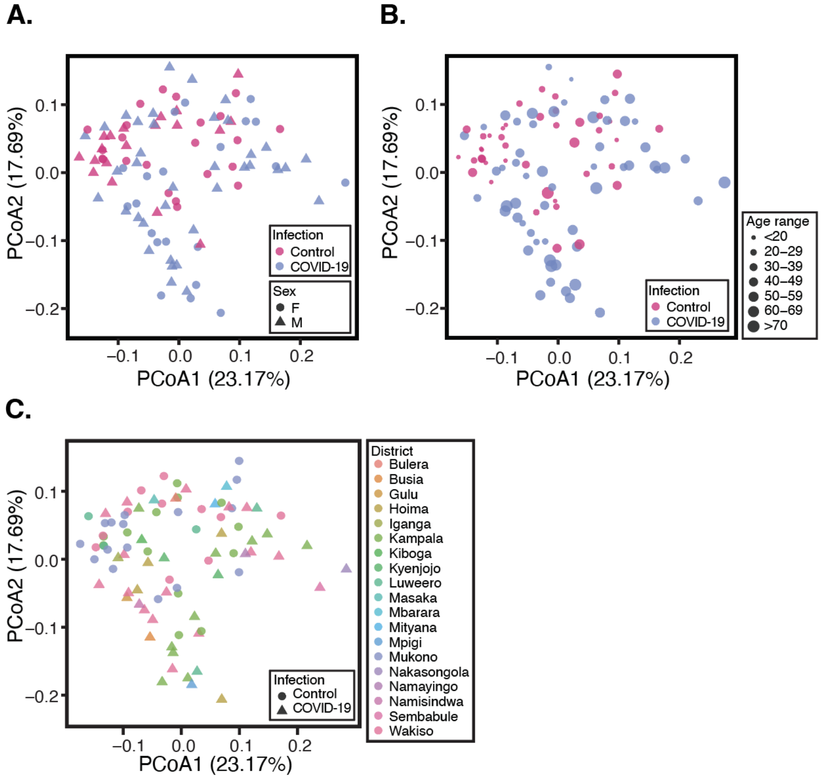


**Supplemental Fig. 1. A.** Principal coordinate analysis (PCoA) calculated using the weighted UniFrac distance metric is shown for males and females (p = 0.374, PERMANOVA; p = 0.622, PERMDISP). The same PCoA is also plotted with point size indicating age range (**B**; p < 0.01, PERMANOVA; p < 0.01, PERMDISP) or point color indicating district of residence (**C**).

#### Suppl. Fig 2.

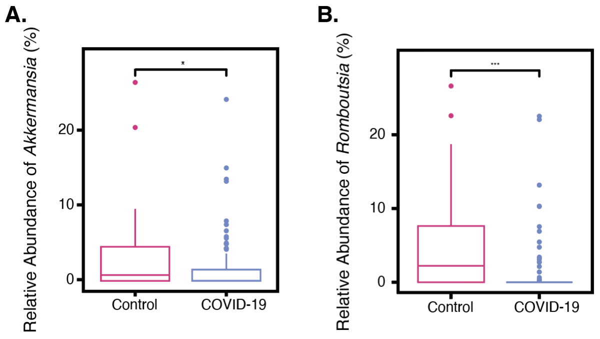


**Supplemental Fig. 2** Genera differentially abundant between COVID-19 cases and controls. The relative abundance of **A** *Akkermansia* species (p* < 0.05, Kruskal-Wallis) and **B** *Romboutsia* species (p*** < 0.0005, Kruskal-Wallis) in positive COVID-19 individuals and asymptomatic individuals.

**Suppl. Fig. 3**

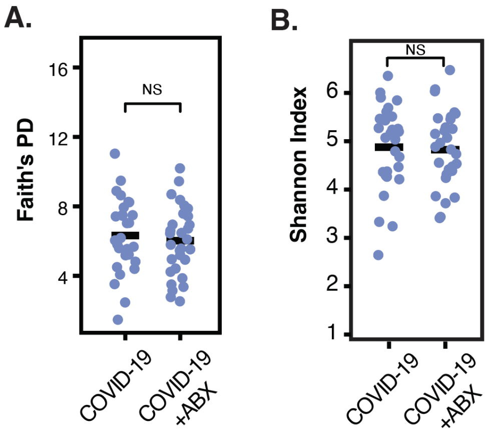


**Supplemental Fig. 3** The gut microbiome of individuals with COVID-19 with antibiotic treatment is not significantly different than individuals with COVID-19 without antibiotics. Alpha diversity is shown using **A** Faith's Phylogenetic Diversity (PD) and **B** the Shannon Index (p = 0.544 Faith's PD, p = 0.644 Shannon) (ABX = antibiotics, not significant - NS, Kruskal-Wallis).

**Suppl. Fig. 4**

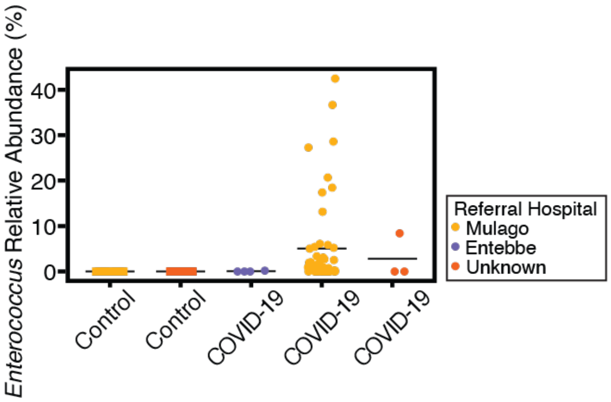


**Supplemental Fig. 4** *Enterococcus* abundance plotted by referral hospital the sample was collected at.
